# Portrait of Mental Health Identified by People with the Post-Covid Syndrome

**DOI:** 10.1101/2023.11.15.23298598

**Authors:** Nancy Mayo, Stanley Hum, Mohamad Matout, Lesley K Fellows, Marie-Josée Brouillette

## Abstract

**Aims:** The Post-COVID-19 syndrome (PCS) represents an epidemic within the COVID-19 pandemic, with potentially serious consequences for affected individuals, the healthcare system, and society at large. Facing a new and poorly understood health condition, this study aimed to produce a patient-centered understanding of mental health symptom patterns, functional impact, and intervention priorities.

**Methods:** A cross-sectional analysis of the first 414 participants in a longitudinal study recruited over a 5- month from September 2022 to January 2023 was carried out involving people from Quebec who self-identified as having symptoms of PCS. People were asked to name areas of their mental health affected by PCS using the structure of the Patient Generated Index (PGI), an individualized measure suited to eliciting the most frequent and most bothersome symptoms. The PGI was supplemented with a set of patient-reported outcome measures across the rubrics of the Wilson- Cleary model. The text threads from the PGI were grouped into topics using BERTopic analysis.

**Results:** Twenty topics were identified from 818 text threads referring to PCS mental health symptoms nominated using the PGI format. 35% of threads were identified as relating to anxiety, discussed in terms of five topics: generalized/social anxiety, fear/worry, post-traumatic stress, panic, and nervous. 29% of threads were identified as relating to low mood, represented by five topics: depression, discouragement, emotional distress, sadness, and loneliness. A cognitive domain (22% of threads) was covered by four topics referring to concentration, memory, brain fog, and mental fatigue. Topics related to frustration, anger, irritability. and mood swings (7%) were considered as one domain and there were separate topics related to motivation, insomnia, and isolation.

**Conclusion:** This novel method of digital transformation of unstructured text data uncovered different ways in which people think about classical mental health domains. This information could be used to evaluate the extent to which existing measures cover the content identified by people with PCS or to justify the development of a new measure of the mental health impact of PCS.

## Introduction

The Post-COVID-19 syndrome (PCS), characterized by a constellation of persistent symptoms following a SARS-CoV-2 infection, has emerged as a significant and challenging health issue in the wake of the global COVID-19 pandemic. While the pandemic itself has brought to light numerous complexities in healthcare and society, PCS represents an epidemic within the pandemic, with potentially serious consequences for affected individuals, the healthcare system, and society.

The prevalence of PCS is very difficult to estimate as there is no established criteria defining the syndrome, except time. Case identification relies on persons self-identifying the persistence of symptoms. Best estimates come from population-based methods. From a random sample of the Canadian population, an estimated 40% were infected with COVID-19 and 14.8% of the Canadian population had symptoms past 3 months consistent with definition of PCS.[1]. The UK included a single question about PCS on the 2021 census and estimated that 3% were affected. [2]

A systematic review of studies (n=76) sampling from those infected with SARS-CoV-2 virus conducted between January 2020 and August 2022 reported that up to 12-months post-infection with nearly 59% (range 14-100) of individuals reported experiencing some form of symptom related to PCS. Among the most common specific symptoms were fatigue, post-exertional malaise, disordered sleep, shortness of breath, anxiety, and depression, affecting anywhere from 32% to 47% of individuals.[3, 4]

Mental health challenges within the PCS population have been frequently documented including anxiety, depression, post-traumatic stress, cognitive impairment, and sleep disturbances. These challenges have been typically assessed through symptom checklists[5], generic measures such as the PHQ-9 and GAD-7, and generic health-related quality of life measures like the EQ-5D.[6–8]

While existing measures have provided valuable insights into the mental health aspects of PCS, it is essential to recognize that PCS is a distinct and relatively novel syndrome that is unlikely to disappear anytime soon. As such, there remains an ongoing need to develop comprehensive measures that capture its unique impact comprehensively so that specific and personalized therapies can become the standard of care.

To address this knowledge gap, the Quebec Action for Post-COVID (QAPC) cohort study was designed. Our research endeavors to capture a broad spectrum of outcomes relevant to PCS, including those traditionally considered indicative of mental health challenges. In this study, we addressed these gaps by adopting a patient-driven approach to mental health assessment in PCS. Rather than relying exclusively on conventional mental health measures, this study employs an individualized measure, the Patient Generated Index (PGI)[9], to gain insights directly from individuals experiencing PCS. Coupled with an advanced Natural Language Processing method, this study aims to provide a more in-depth understanding of the key mental health issues identified by people experiencing PCS. This innovative combination allows us to analyze self-reported symptoms in a structured manner, mapping them onto key mental health topics. At the time of enrollment in the study, the COVID vaccine was widely available and most of the restrictions on social contacts had been lifted.

## Methods

This is a planned analysis of participants in the Quebec Action for Post-COVID cohort. QAPC is a longitudinal study of Quebec residents self-identifying with persistence of COVID-19 symptoms for more than 12 weeks after a confirmed or presumed infection with SARS-Cov2 virus. Enrollment was from September 2022 to September 2023 (13 months) with ongoing assessments every 3 months. A cross-sectional analysis of the first 414 QAPC participants recruited over the first 5 months was carried out for this paper. This longitudinal study, launched in September 2022, focuses on residents of Quebec, Canada, who self-identify as experiencing PCS symptoms lasting more than 12 weeks following a confirmed or presumed SARS-CoV-2 infection. Assessments every three months are ongoing. A full description of the functional consequences of PCS has been previously published [10].

The project (2022-8066) was approved by the Research Ethics Board of the McGill University Health Centre. Following an e-consent process, participants responded to the study questionnaires on the web-portal “Research Electronic Data Capture” (REDCap).

### Measurement

The overall measurement framework QAPC this study was the World Health Organization’s International Classification of Functioning, Disability and Health (ICF) [11] with an extension to health-related quality of life and quality of life based on the Wilson-Cleary Model[12].

For the measurement of mental health, the brain health framework as defined by Chen et al. [13] was used: “a lifelong dynamic state of cognitive, emotional, and motor domains underpinned by physiological processes. It is multidimensional and can be objectively measured and subjectively experienced”. Mental health symptoms were collected using the Visual Analogue Health States (VAHS)[14] for distress and sleep, the RAND-36 Mental Health Index[15], and a screen for post-traumatic stress disorder (PTSD)[16]. Associated outcomes related to cognitive concerns[17], loneliness, and irritability were also collected. Information on motor domains is not reported here but physical function is described in an earlier paper .[10]

Data on participants’ experiences with mental health symptoms was collected using a modification of the Patient Generated Index (PGI)[9]. In the original version, there are three steps. First, respondents are asked to name five areas where their health condition has affected their life. Second, each area is then rated on a scale from 0 to 10 where 0 is “the worst you could imagine” and 10 is “exactly as you would like to be”. For the third step, the person distributes 10 imaginary spending points across the 5 areas according to which areas they would most like to see improved. When only five health areas are asked for, the PGI would best represent the construct of health-related quality of life. There is also a version with a 6^th^ area that is for non-health related areas (which are not specified) and then 12 spending points are distributed. This version would represent the construct of quality of life. This process avoids the over-reporting that can occur when people are asked to choose from a list of symptoms, rather than spontaneously declaring them[18], and provides information about priorities in the person’s own words.

For eliciting mental health areas of concern, Step 1 of the PGI, participants were instructed as follows: *“After COVID, people can have challenges to their mental health such as feelings of anxiety, depression or emotional distress. We would like you to think about the most bothersome symptoms of your mental health. Please list up to 5 symptoms that are the most bothersome for you. Please list symptoms here even if you mentioned them previously.*“ Step 2 was modified to made 10 the “worst imaginable severity” in keeping with eliciting negative effects of PCS. Step 3 was identical to the original PGI.

Text threads were gathered in both English and French. Googletrans 3.1.0a0 python library that implemented Google Translate API was used to translate French text to English before further analyses.[19]

### Statistical Methods

A total score for the PGI was calculated by multiplying each area’s rating from 0 to 10 by the number of spending points allocated and summing. This yields a score from 0 to 100 with 100 representing the worst mental health imaginable. To compare with other mental health measures, the value was converted to have 100 as the best mental health.

Sentence Bidirectional Encoder Representations from Transformers (SBERT) model used to extract topics from the text threads. Topic modeling is a common Natural Language Processing (NLP) task used to uncover abstract themes (topics) within unstructured text. Bertopic[20], a topic modeling method based on embeddings from pre-trained sentence transformers (all-MiniLM-L6- v2) was implemented. This allows the semantic context of the sentence/text to be considered when extracting topics and not just based on lexicon similarities.

Uniform Manifold Approximation and Projection (UMAP)[21] algorithm was used to reduce dimensionality of the embeddings, and hierarchical density-based spatial clustering (HDBSCAN)[22] was applied to identify dense clusters in the documents. Model parameters were set to reduce outliers and remove stop words. CountVectorizer from scikit-learn[23] after the embedding and clusters were generated to remove stop words (e.g. “the”, “a”, “an”, “so”, etc…) to make the topics more readable. Class-based term frequency inverse document frequency (c-TF-IDF) was used to compare the importance of the terms in each cluster.[24]. UMAP random state was set to 42 to allow the model to be reproducible. Bertopic does not require the number of topics to be predefined. The large number of topics generated were reduced by combining similar topics based on expert review and hierarchical clustering of embeddings.[20]

The hardware was a Lenovo i5-4570 3.20GHz with 16g DDR3 with a Nvidia RTX3090 GPU. Software environment used was on a Windows 10 Enterprise 64-bit Version 22H2, conda 4.11.0, Python 3.9.16, CUDA 11.8, Jupyter Notebook 6.5.2, bertopic 0.13.0, PyTorch 2.1.0.dev20230408+cu118, and Scipy 1.10.0.

## Results

The study sample comprised 314 women (75.8%), 94 men (22.7%), and 6 identifying as other. The mean age of the women was 48.2 years (SD: 11.6) and of the men, 51.6 years (SD: 13.1). Table 1 presents selected characteristics of the mental health of the study sample. Participants scored, on average, 53.6 out of 100 on the PGI-Mental Health (higher is better). This is very close to their score on the Mental Health Index from the RAND-36 which was 54.0 out of 100 much lower than the normative value of 75.6 in the general population of this age range. The sample also reported poor cognition, with a score of 43.5 out of 100 on a measure of self-reported memory and attention abilities, norm 81 (SD: 20)[17].

**Table 1:** Mental Health Characteristics of the QAPC Study Sample (n=414)

| **Symptom [Norm]** | **N or Mean [Median]** | **% or SD** |
| --- | --- | --- |
| *Measures with 100 as best* |  |  |
| PGI-Mental Health | 53.6 [40.0] | 35.1 |
| Mental Health Index [75.6<sup>a</sup>] | 54.0 [56.0] | 20.9 |
| Cognition:C3Q (0-100 best) [81<sup>b</sup>] | 43.5 [41.6] | 26.4 |
| *Measures with high values as worst* |  |  |
| Lonely sometimes or often [10.4%<sup>c</sup>] | 254 | 61.4% |
| PTSD ($\geq 3/5$ symptoms) | 103 | 24.9% |
| Irritable often to always | 88 | 21.2% |
| Distress (VAS 0 – 100) | 38.8 [36.5] | 29.9 |
| Sleep (VAS 0 -100) | 50.0 [54.5] | 29.6 |

<sup>a</sup> Population rate of loneliness are from Statistics Canada
<sup>b</sup> Norms for C3Q are from Askari et al.[17]
<sup>c</sup> Population rate of loneliness are from Statistics Canada [30]

For measures on negative mental health constructs, 61.4% indicated that they were sometimes or often lonely, compared with 10.4% of the general public in the pre-COVID period; 24.9% met criteria for PTSD, and 21.2% reported they were often to always irritable. Distress rating was 38.8 and sleep rating was 50 where 100 is the worst imaginable rating.

Figure 1 shows the 20 topics emerging from the 818 text threads collected using the PGI-Mental Health from the 414 participants. The informative words per topic are visualized in as bars based on the combination of the frequency of the words nominated and the importance of the words within the cluster. The x-axis (c-TF-IDF) provides an indication of the contribution of each word to the topic. Figure 2 summarizes the prevalence of each topic.

**Figure 1.**
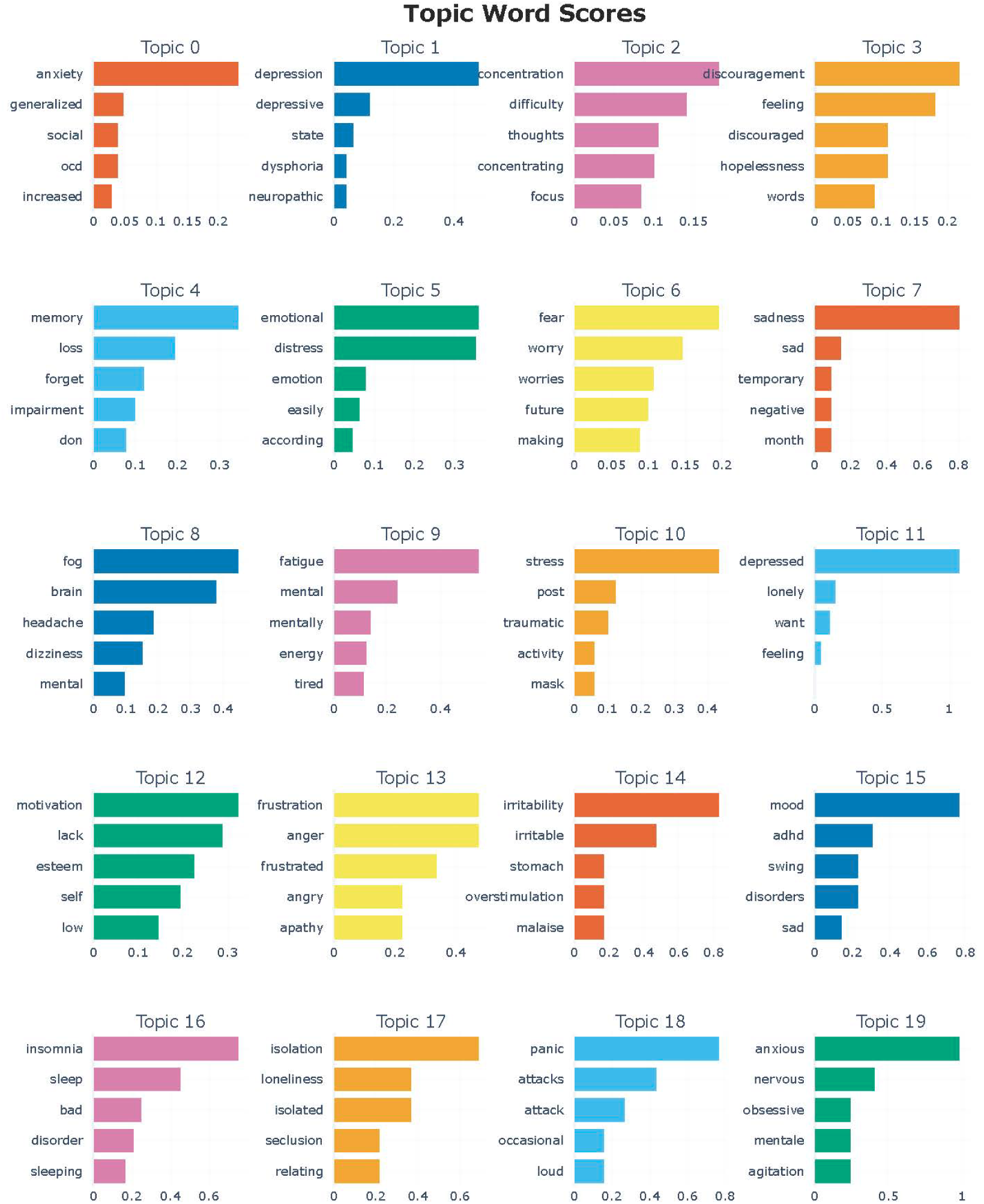
Topics emerging from the 818 text threads people with PCS used to describe their mental health symptoms using the Patient Generated Index.

**Figure 2.**
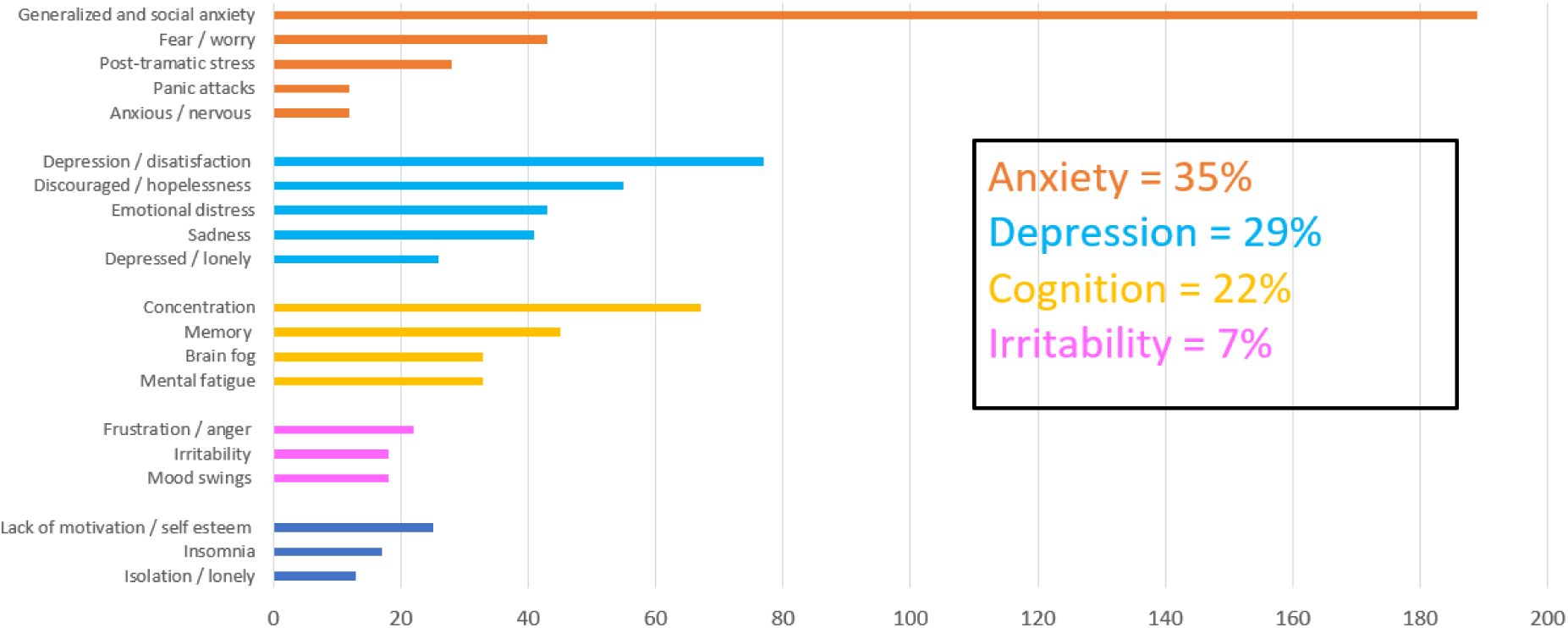
Prevalence of topics related to anxiety, depression, cognition, irritability, and others. *56 of the 818 text threads were not classified into topics

The most common topic (Topic 0) was related to anxiety, specifically anxiety associated with the words generalized and social. This was the most prevalent topic with over 189 nominations (23.1%). However, anxiety was included in other topics associated with fear and worry (Topic 6), post-traumatic stress (Topic 10), panic attacks (Topic 18), and being nervous (Topic 19). Overall participants, one of these forms of anxiety were identified by 34.2% of participants. Similarly, depression was represented by five different topics: depression/dysphoria (Topic 1); depression/discouraged (Topic 3); depression/emotional distress (Topic 5); depression/sadness (Topic 7), and depression/lonely (Topic 11). Topics related to depression were nominated by 29% of participants. Cognition, nominated by 22% of the sample, was represented by topics related to concentration, memory, brain fog, and mental fatigue (Topics 2, 4, 8 and 9). People also expressed frustration/anger, irritability, and mood swings (Topics 13, 14, and 15) as well as lack of motivation, insomnia and isolation (Topics 12, 15, and 17) with lower frequency.

## Discussion

The primary objective of this research was to gain insight into the experiences of individuals with Post-Covid Syndrome (PCS) from a patient-centered perspective, focusing on symptom patterns, functional consequences, and intervention priorities. The study identified twenty distinct mental health topics, with anxiety emerging as the most prevalent concern.

Our findings indicate that anxiety is a major apprehension for individuals dealing with PCS, with 35% of textual discussions centered around anxiety. This finding aligns with prior research acknowledging anxiety as a common symptom of PCS.[25, 26]. Our study also identified five subtopics within anxiety, encompassing generalized/social anxiety, fear/worry, post-traumatic stress, panic, and nervousness. These subtopics offer a more precise insight into the various facets of anxiety in PCS individuals. Post-traumatic stress disorder was a topic likely because a proportion of the sample (11%) identified stressful events were witnessed as part of their job.

Another prevalent symptom was low mood, affecting 29% of the textual threads, encompassing depression, discouragement, emotional distress, sadness, and loneliness. This corresponds with earlier research identifying depression as a common feature of PCS.[26, 27] Our study revealed four subtopics associated with cognitive domains, including concentration, memory, brain fog, and mental fatigue, hinting at potential cognitive impairments affecting daily functioning in PCS individuals. Cognitive effects of PCS are frequently documented.[28] While cognitive difficulties are a common symptom of depression[29], here they were never in the same topic suggesting two different phenomena.

This study provides a novel way to assessing mental health difficulties by allowing participants to nominate their most distressing symptoms using the Patient Generated Index (PGI), emphasizing the importance of patient-centered perspectives. This approach permits individuals to express their experiences in their own words, thus minimizing the risk of over-reporting and providing a more holistic comprehension of their distinctive challenges.

The application of BERTopic analysis, an advanced natural language processing technique, played a pivotal role in converting unstructured text data into meaningful topics. This method afforded us a deeper understanding of how individuals with PCS conceptualize and classify their mental health experiences, offering valuable insights for customizing interventions and support strategies.

However, it is crucial to acknowledge some limitations in our study. The cross-sectional analysis of the initial 414 participants represents a preliminary exploration of mental health challenges associated with PCS. A longitudinal perspective could offer valuable insights into the evolution of these challenges over time. Our sample self-identified with PCS and will not represent all persons who have experienced PCS. Nevertheless, the sample would represent people seeking solutions to PCS through the health care system. There is no standard way of identifying people with PCS short of a national survey such as was conducted in Canada and Britain.[1, 2] However, these population based approaches are not suitable for obtaining a comprehensive assessment of the effects of PCS. It is also possible to obtain information about people with PCS by following those from a standard sampling frame such as a hospitalized population but in Canada only 6% of the population were hospitalized with an estimated 40% infected.[1]

The diversity of the mental health topics raised by people with PCS should be helpful to clinicians helping people with PCS to recover. Future research endeavors should delve into the underlying factors contributing to these mental health concerns, including potential biological, psychological, and social determinants. Longitudinal studies could provide clarity on the trajectories of these mental health challenges and their connections to other clinical outcomes.

## Data Availability

The data comprise text and may be made available upon reasonable request to authoer

## References

1. Statistics-Canada. Long-term symptoms in Canadian adults who tested positive for COVID-19 or suspected an infection, January 2020 to August 2022. Available from: https://www150.statcan.gc.ca/n1/daily-quotidien/221017/dq221017b-eng.htm.

2. Census, U., Prevalence of ongoing symptoms following coronavirus (COVID-19) infection in the UK: 7 July 2022. 2021.

3. Kuodi, P., Y. Gorelik, and M. Edelstein, Characterisation of the long-term physical and mental health consequences of SARS-CoV-2 infection: A systematic review and meta-analysis protocol. PLoS One, 2022. 17(4): p. e0266232.

4. Kuodi, P., et al., Characterization of post-COVID syndromes by symptom cluster and time period up to 12 months post-infection: A systematic review and meta-analysis. Int J Infect Dis, 2023. 134: p. 1–7.

5. Benoit-Piau, J., et al., Long-Term Consequences of COVID-19 in Predominantly Immunonaive Patients: A Canadian Prospective Population-Based Study. J Clin Med, 2023. 12(18).

6. Guillen-Burgos, H.F., et al., Factors associated with mental health outcomes after COVID-19: A 24-month follow-up longitudinal study. Gen Hosp Psychiatry, 2023. 84: p. 241–249.

7. Ashton Rennison, V.L., C.J. Chovaz, and S. Zirul, Cognition and psychological well-being in adults with post COVID-19 condition and analyses of symptom sequelae. Clin Neuropsychol, 2023: p. 1–28.

8. Sahanic, S., et al., COVID-19 and its continuing burden after 12 months: a longitudinal observational prospective multicentre trial. ERJ Open Res, 2023. 9(2).

9. Ruta, D.A., et al., A new approach to the measurement of quality of life. The Patient-Generated Index. Medical Care, 1994. 32(11): p. 1109–1126.

10. Mayo, N., et al., A patient-centered view of symptoms, functional impact, and priorities in post-COVID-19 syndrome: Cross-sectional results from the Québec Action Post-COVID cohort. medRxiv, 2023: p. 2023.05.27.23290638.

11. Who, World Health Organization. International Classification of Functioning, Disability and Health. Vol. Second revision. 2001, Geneva.

12. Wilson, I.B. and P.D. Cleary, Linking clinical variables with health-related quality of life. A conceptual model of patient outcomes. Journal of the American Medical Association, 1995. 273(1): p. 59–65.

13. Chen, C., et al., Global Prevalence of Post-Coronavirus Disease 2019 (COVID-19) Condition or Long COVID: A Meta-Analysis and Systematic Review The Journal of Infectious Diseases, 2022. 226(9): p. 1593–1607.

14. Rosenzveig, A., et al., Toward patient-centered care: a systematic review of how to ask questions that matter to patients. Medicine (Baltimore), 2014. 93(22): p. e120.

15. Hays, R.D., C.D. Sherbourne, and R.M. Mazel, The RAND 36-Item Health Survey 1.0. Health Econ., 1993. 2(3): p. 217–227.

16. Prins, A., et al., The Primary Care PTSD Screen for DSM-5 (PC-PTSD-5): Development and Evaluation Within a Veteran Primary Care Sample. J Gen Intern Med, 2016. 31(10): p. 1206–11.

17. Askari, S., et al., Development and validation of a voice-of-the-patient measure of cognitive concerns experienced by people living with HIV. Qual Life Res, 2021. 30(3): p. 921–930.

18. Villemure, R., P. Nolin, and N. Le Sage, Self-reported symptoms during post-mild traumatic brain injury in acute phase: influence of interviewing method. Brain Inj, 2011. 25(1): p. 53–64.

19. Han, S. googletrans 3.0.0a0. 2020; Available from: https://pypi.org/project/googletrans.

20. Grootendorst, M., BERTopic: Neural topic modeling with a class-based TF-IDF procedure. arXiv, 2022.

21. McInnes, L., et al., UMAP: Uniform Manifold Approximation and Projection. Journal of Open Source Software, 2018. 3(29).

22. McInnes, L., J. Healy, and S. Astels, hdbscan: Hierarchical density based clustering. The Journal of Open Source Software, 2017. 2(11).

23. Pedregosa, F., et al., Scikit-learn: Machine Learning in Python. Journal of Machine Learning Research, 2012. 12.

24. Sánchez-Franco, M.J. and M. Rey-Moreno, Do travelers’ reviews depend on the destination? An analysis in coastal and urban peer-to-peer lodgings. Psychology & Marketing, 2021. 39(2): p. 441–459.

25. Mazza, M.G., et al., One-year mental health outcomes in a cohort of COVID-19 survivors. J Psychiatr Res, 2021. 145: p. 118–124.

26. Taquet, M., et al., Incidence, co-occurrence, and evolution of long-COVID features: A 6- month retrospective cohort study of 273,618 survivors of COVID-19. PLoS Med, 2021. 18(9): p. e1003773.

27. Mazza, M.G., et al., Prevalence of depression in SARS-CoV-2 infected patients: An umbrella review of meta-analyses. Gen Hosp Psychiatry, 2023. 80: p. 17–25.

28. Ceban, F., et al., Fatigue and cognitive impairment in Post-COVID-19 Syndrome: A systematic review and meta-analysis. Brain Behav Immun, 2022. 101: p. 93–135.

29. Perini, G., et al., Cognitive impairment in depression: recent advances and novel treatments. Neuropsychiatr Dis Treat, 2019. 15: p. 1249–1258.

30. Statistics-Canada. Available from: https://www150.statcan.gc.ca/n1/daily-quotidien/211124/dq211124e-eng.htm.

